# FusionAge framework for multimodal machine learning-based aging clocks uncovers cardiorespiratory fitness as a major driver of aging and inflammatory drivers of aging in response to spaceflight

**DOI:** 10.1101/2025.11.27.25341167

**Authors:** Robert Chen, Nicholas Bartelo, Mohith Arikatla, Christopher E. Mason, Olivier Elemento

## Abstract

Traditional epigenetic aging clocks are limited because they do not incorporate clinical information and functional tests, and rely on DNA samples and methylation profiling infrastructure which are not easily accessible. To address these limitations, we built a new framework, FusionAge, with which we trained 26 aging clocks using interpretable nonlinear models, including deep neural networks (DNNs). Our results show that multimodal clocks built with DNNs significantly outperform clocks derived from single modalities or traditional linear models. FusionAge-derived biological age is more strongly associated with incident disease and mortality compared to chronological age in UK Biobank individuals. We validated these findings in the National Health and Nutrition Examination Survey, confirming that cardiorespiratory fitness is a major, consistent driver of biological age. Finally, we applied FusionAge to demonstrate its utility in detecting biological age changes in astronauts following spaceflight. Together, we demonstrate a powerful, portable framework for assessing biological age that captures the complex, multifactorial nature of human aging.

## Introduction

Accurate aging biomarkers are essential for profiling healthy individuals longitudinally, interpreting personalized drivers of biological age on the individual level, predicting disease risk and evaluating the impacts of activities and other stressors sustained by the human body.

Previously reported aging clocks have been developed to predict chronological age using various types of data, including DNA methylation, gene expression, and clinical biomarkers, and a predominance of them are based on epigenetic markers^1–9^. However, since epigenetic features are not traditionally collected in clinical settings, such aging clocks have limited potential to be scored on individuals in typical healthcare settings. Furthermore, a majority of these aging clocks use variations of linear regression which do not allow for the model to capture nonlinear relationships between features. Additionally, a limitation of epigenetic clocks is the interpretability, as many individual epigenetic islands in isolation do not have obvious disease associations.

Various works have proposed using other structured features such as proteomics, metabolomics, glycomics, clinical data and movement sensor data^10,11^. Metabolomics based clocks have been proposed, which typically involve analytes measured via mass spectrometry ^12–15^. Various studies have linked metabolomic age with aging-related disease and mortality ^16^. Similarly, proteomics based clocks using plasma protein abundances have been proposed ^17,18^. While metabolomic and proteomic clocks in isolation can be associated with disease incidence and mortality, common drawbacks include noisy data (including measurements that are easily obscured by acute changes in diet), as well as lack of interpretability when used in isolation. Various works have proposed using clinical data^3,19–21^, which are commonly interpreted in a medical management setting but in isolation they do not capture in depth functional abilities of the individual.

Multimodal machine learning based approaches which combine datasets across modalities ^22,23^ (e.g., clinical, proteomic, metabolomic, imaging, functional tests), have been proposed as potential strategies to integrate distinct data types for challenging tasks such as clinical outcome prediction in COVID-19 ^24,25^, patient stratification^26^, remote patient monitoring for applications such as fall detection^27^, digital twins for clinical trials and drug discovery ^28,29^.

Regarding aging clocks, Tian et al 2023 ^30^ utilized manually curated subsets of clinical data and imaging related features to predict organ-specific ages from individuals in the UK Biobank (UKB) cohort. However, they did not incorporate several key modalities which have previously been demonstrated to be strong predictors of age-related morbidity, such as cardiorespiratory fitness ^31^, and strong predictors of age and organ-specific aging-related disease, such as proteomics ^17,18^. Various methods separate features into organ-specific subsets to compute “organ ages”^18,30^; such scores for biological age may carry associations with specific diseases, but a limitation is that many human diseases are not organ-specific by nature, and involve a complex interplay between physiological and molecular drivers, and thus aging should not be assessed via organ-specific domains, nor should biological aging be assessed by single domains of information.

To address these gaps, we developed FusionAge, a multimodal machine learning-based clock training framework that facilitates training of both modality-specific and multimodal aging clocks using a variety of data types: biospecimens, physical measures, functional tests and biomedical imaging. FusionAge addresses various shortcomings of previous aging clocks:

1. **Multimodal Architecture:** Many aging clocks use features from a limited number of domains (e.g., epigenetic data only). FusionAge is designed to integrate diverse data types—including imaging and functional tests common in healthcare settings—and explicitly tests different fusion strategies to create a more holistic model of aging.
2. **Data-Driven Feature Modalities**: Rather than pre-selecting features based on organ systems, which may miss systemic or multi-organ disease drivers, FusionAge groups features by their data collection modality. This data-driven approach allows for the unbiased discovery of the most predictive features for disease and mortality.
3. **Nonlinear Modeling**: The majority of aging clocks rely on linear models 1–3. Our method employs a thorough model selection process that includes nonlinear models like XGBoost and deep neural networks, which can capture complex, non-additive relationships between features and age.
4. **Portability and Interpretation**: By using data modalities common in clinical settings, our framework is highly portable to new datasets. Furthermore, the framework includes an interpretability module that allows for an efficient understanding of the most contributory features to biological age, on both the population and individual level, which we apply to generate hypotheses from our models.

We applied the FusionAge framework to first train 22 modality-specific clocks to establish performance benchmarks for individual data types. We then tested the central hypothesis that combining data from multiple domains into multimodal clocks would improve age prediction and provide deeper biological insights than any single modality alone. To this end, we constructed 4 multimodal clocks using both early and late fusion architectures and a systematic search of various regression algorithms. We assessed each clock’s performance via standard metrics (Pearson correlation, mean absolute error) and its association with disease and mortality.

As external validation of the methodology, we trained multimodal aging clocks with the FusionAge framework in the CDC National Health and Nutrition Examination Survey (NHANES)^32^ biobank, which validated cardiorespiratory fitness as a major driver of biological aging. Finally, we scored a deep neural network based aging clock trained with the FusionAge framework on a cohort of astronauts in the Inspiration4 space mission, which uncovers putative pathways representing inflammatory changes and tissue remodeling in response to space travel.

## Results

### FusionAge strongly predicts biological age

We proposed the FusionAge framework for training multimodal aging clocks and applied it to train 26 aging clocks on a training dataset from the UK Biobank (UKB) consisting of 502,366 individuals aged 37 to 73 years (229,068 males and 272,298 females). Aging clocks were trained on all major domains of features in the UK Biobank including physical measures (anthropometric features, arterial stiffness, bone density, body impedance, vital signs), biospecimens (blood chemistry, urine chemistry, metabolomics, proteomics, telomere characteristics), functional tests (cognitive test, eye measures, cardiorespiratory fitness, skeletal muscle strength, hearting test, spirometry) and imaging (abdominal MRI, brain MRI, carotid ultrasound, DEXA scan, electrocardiogram, heart MRI) (Fig. 1a-c, Table 1). Aging clocks are defined as models that utilize features in a regression against chronological age (CA), resulting in an output prediction of biological age (BA). Aging clocks are trained via a multimodal architecture utilizing either early fusion or late fusion on a specified feature set (Table 1), combined with a specific regression algorithm (referred to in the format “FusionAge-<ALGORITHM>-<FEATURE SET>”; regression algorithms included linear regression (LR), lasso, ElasticNet, XGBoost and deep neural network (DNN)). A total of 22 modality-specific aging clocks and 4 multimodal aging clocks were trained and assessed on the UKB training dataset (Methods).

**Fig. 1:**
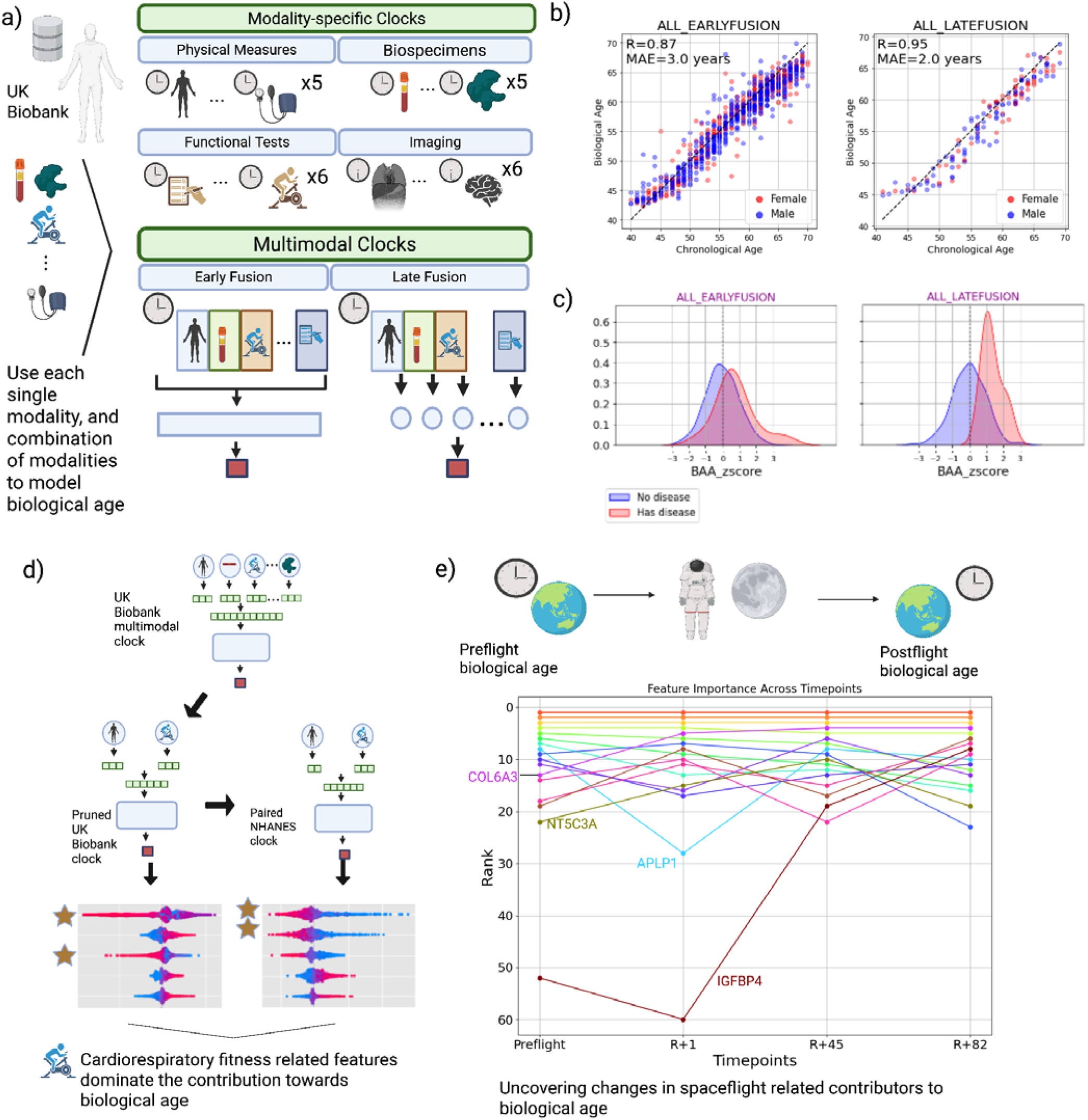
FusionAge multimodal clocks strongly predict biological age and are readily applied to clinical data-rich datasets. **a)** general overview of FusionAge clock training framework. Data are extracted and mapped to measurement modalities; a total of 22 modality-specific clocks and 4 multimodal clocks are trained with the framework. **b)** multimodal clocks strongly predict biological age, with a Pearson correlation coefficient of R=0.95 for a late fusion clock based on physical measures, biospecimens, and functional tests and R=0.91 for a late fusion clock based on imaging data. **c)** multimodal clocks are strongly correlated with aging-related disease. **d)** paired multimodal clocks trained on UK Biobank and NHANES cohorts retain strong performance and validate cardiorespiratory fitness as the most contributory driver to biological age. **e)** demonstration of FusionAge towards a cohort of astronauts from the Inspiration4 space mission demonstrates alternating ranking of contributions of individual features towards biological age.

**Table 1:** all clocks trained, labeled by modality feature set.

| Modality Feature Set Abbreviation | Modality Description |
| --- | --- |
| **Modality-specific clocks** |  |
| ANTHROPOMETRIC | Anthropometric features |
| ARTERIALSTIFFNESS | Arterial stiffness |
| BONEDENSITY | Bone density |
| CHEM-BLOOD | Blood chemistry |
| CHEM-URINE | Urine chemistry |
| COGNITIVE | Cognitive test |
| EYE | Eye measures |
| FITNESS-CARDIO | Cardiorespiratory fitness |
| FITNESS-STRENGTH | Skeletal muscle strength |
| HEARING | Hearing test |
| IMPEDANCE | Body impedance |
| METABOLOMICS | Metabolomics |
| PROTEOMICS | Proteomics |
| SPIROMETRY | Spirometry |
| TELOMERE | Telomere |
| VITALSIGNS | Vital signs |
| ABDOMINAL-MRI | Abdominal MRI |
| BRAIN-MRI | Brain MRI |
| CAROTID-ULTRASOUND | Carotid ultrasound |
| DEXA | DEXA scan |
| ECG | ECG |
| HEART-MRI | Heart MRI |
| **Multimodal clocks** |  |
| ALL_EARLYFUSION | **Early fusion:** anthropometric features, arterial stiffness, bone density, blood chemistry, urine chemistry, cognitive test, eye measures, cardiorespiratory fitness test, strength test, hearing test, body impedance, metabolomics, proteomics, spirometry test, telomeres and vital signs |
| ALL_LATEFUSION | **Late fusion:** anthropometric features, arterial stiffness, bone density, blood chemistry, urine chemistry, cognitive test, eye measures, cardiorespiratory fitness test, strength test, hearing test, body impedance, metabolomics, proteomics, spirometry test, telomeres and vital signs |
| IMAGING_EARLYFUSION | **Early fusion:** abdominal MRI, brain MRI, carotid ultrasound, DEXA scan, electrocardiogram and heart MRI |
| IMAGING_LATEFUSION | **Late fusion:** abdominal MRI, brain MRI, carotid ultrasound, DEXA scan, electrocardiogram and heart MRI |
| **Multimodal clocks - External Validation on NHANES dataset** |  |
| ANTHROPOMETRIC | Anthropometric features |
| CHEM-BLOOD | Blood chemistry |
| CHEM-URINE | Urine chemistry |
| DEXA | DEXA scan |
| EYE | Eye measures |
| FITNESS-CARDIO | Cardiorespiratory fitness |
| FITNESS-STRENGTH | Skeletal muscle strength |
| IMPEDANCE | Body impedance |
| SPIROMETRY | Spirometry |
| TELOMERE | Telomere |
| VITALSIGNS | Vital signs |
| NHANES_EARLYFUSION | **Early fusion:** blood chemistry, urine chemistry, eye measures, cardiorespiratory fitness, body impedance |
| NHANES_LATEFUSION | **Late fusion:** blood chemistry, urine chemistry, eye measures, cardiorespiratory fitness, body impedance |
| **Multimodal clocks - Spaceflight** |  |
| SPACE_EARLYFUSION | Blood chemistry, proteomics |

The validation dataset was extracted from the US National Health and Nutrition Examination Survey (NHANES) consisting of 101,316 individuals (49,893 males and 51,423 females) with data for feature domains that overlap with those present in the training dataset (anthropometric features, vital signs, blood chemistry, urine chemistry, telomere characteristics, eye measures, cardiorespiratory fitness, skeletal muscle strength, spirometry, body impedance, DEXA scan). We trained 11 modality-specific and 2 multimodal aging clocks on the NHANES dataset; furthermore, we trained a version of the early fusion aging clock on the UKB and scored it on the NHANES dataset to predict biological age. Across the UKB and NHANES datasets, we used holdout test sets to assess FusionAge-derived biological age via performance metrics, incident disease and mortality associations, and interpretability analysis of the drivers of biological age. We applied the approach to the Inspiration4 dataset as a prototype demonstration for longitudinal profiling of biological age and discovery of putative drivers of biological age changes in response to spaceflight.

FusionAge-derived biological age was highly correlated with chronological age (Fig. 1b, 2a); FusionAge-DNN-IMPEDANCE was the modality-specific clock that was most predictive of biological age (Pearson correlation (R) = 0.99, mean absolute error (MAE) = 0.9, root mean squared error (RMSE) = 1.3), followed by the FusionAge-DNN-PROTEOMICS clock (R=0.92, MAE=2.5, RMSE=3.2). Amongst the imaging-based clocks, FusionAge-DNN-BRAIN-MRI was most predictive of biological age, (R=0.84, MAE=3.8, RMSE=4.8), followed by FusionAge-DNN-DEXA (R=0.73, MAE=3.1, RMSE=5.3).

Of note, all FusionAge multimodal clocks, using both early fusion (FusionAge-DNN-ALL_EARLYFUSION, R=0.87, MAE 3.0, RSE 3.8) and late fusion (FusionAge-DNN-ALL_LATEFUSION, R=0.95, MAE=2.0, RMSE=2.4), architectures, outperformed other implementations of clocks from the literature which utilize clinical, imaging or proteomics data - PhenoAge^3^ (R=0.75, MAE=5.0, RMSE=7.4), YeTianSVM^30^ (R=0.66, MAE=4.6, RMSE=5.9), and OrganAge^18,30^ (R=0.81, MAE=5.2, RMSE=6.2) with respect to Pearson correlation coefficient, MAE and RMSE (Table 2).

**Table 2:**
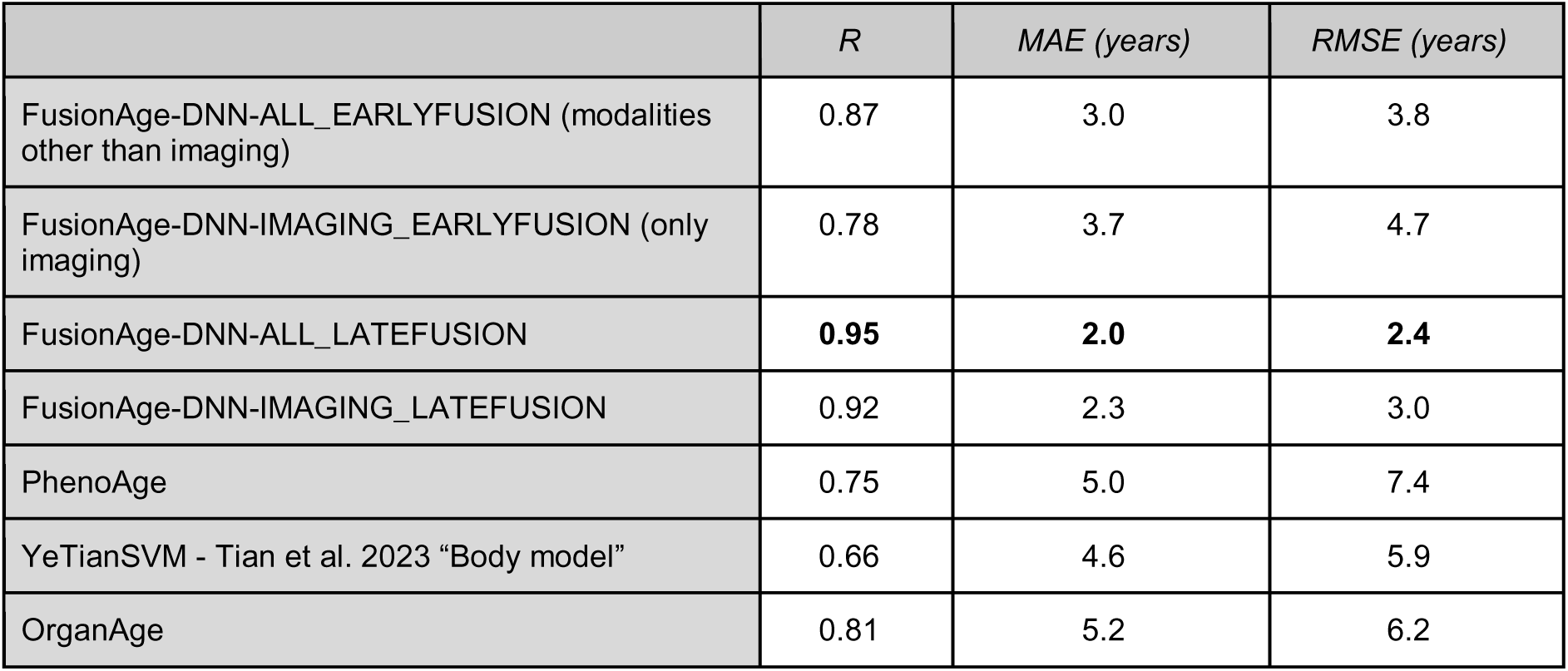
FusionAge performance against baselines. The strongest performance across each metric is indicated in bold font.

Across all DNN-based clocks trained with the FusionAge framework, the pairs of modalities most correlated with each other with respect to biological age are PROTEOMICS with IMPEDANCE (R=0.9), BRAIN-MRI with IMPEDANCE (R=0.9), and BRAIN-MRI with PROTEOMICS (R=0.9) (Fig. 2b). Generally, as the number of features involved in the clock increases, the performance increases. However, modalities such as FITNESS-CARDIO and IMPEDANCE yield strong performance despite low dimensionality of features. Additionally, the multimodal models using feature sets ALL_LATEFUSION and IMAGING_LATEFUSION have strong performance as they use latent representations of multiple clocks’ features which encapsulate knowledge of biological age. Generally, the clocks that were trained on larger training sets had poorer performance; however, this trend is likely due to these data being more easily acquired on a large scale. Multimodal clocks were typically trained on smaller training sets because inclusion of individuals into the training sets is dependent upon having features for all modalities included in the clocks. Despite smaller training set size, performance is greater in these multimodal clocks, which can be attributed to more useful latent representations being encapsulated by these clocks.

**Fig. 2:**
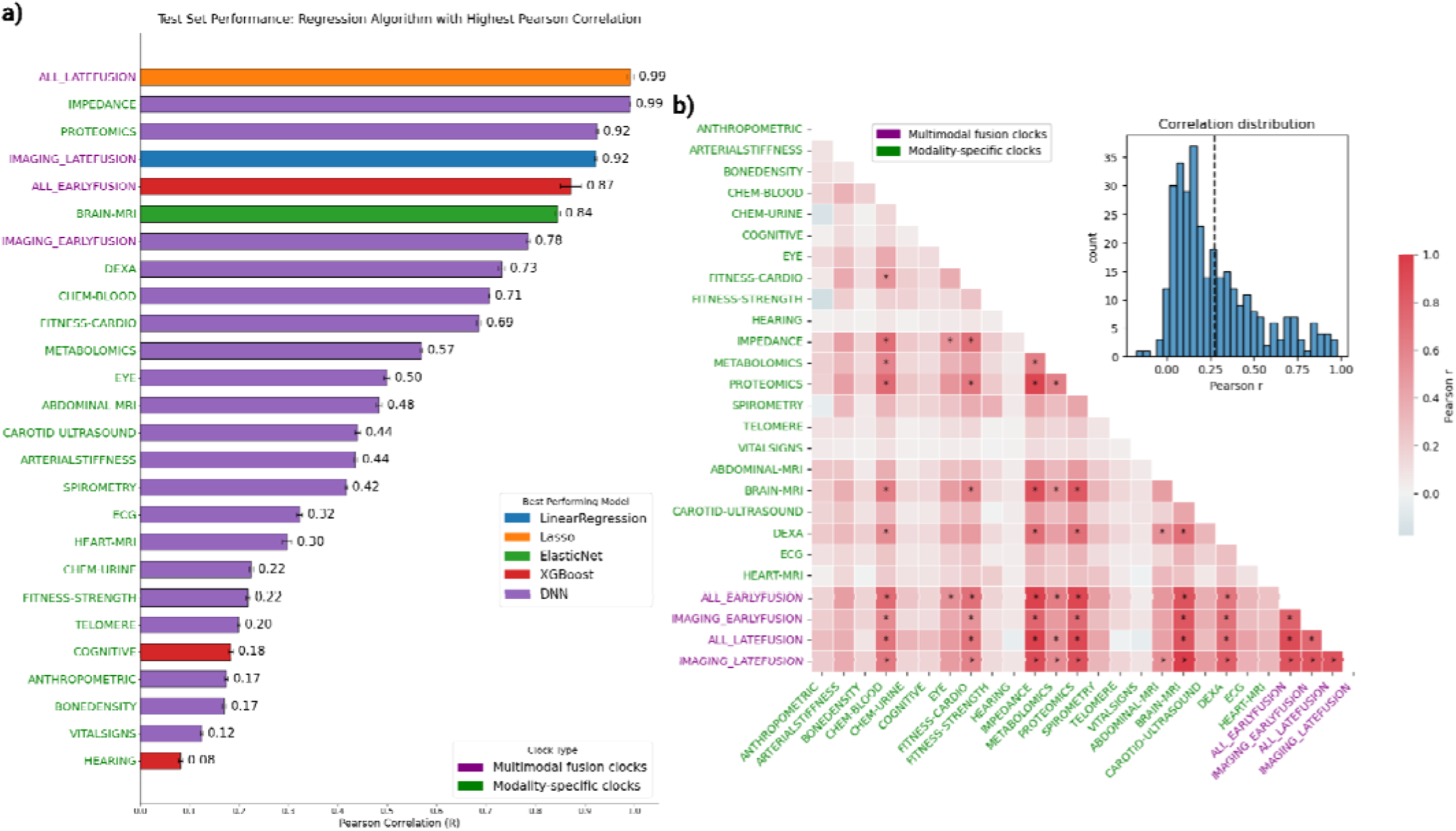
FusionAge-trained modality-specific and multimodal clocks strongly predict biological age across various linear and nonlinear regression algorithms. **a)** performance of modality-specific performance and multimodal clocks; **b)** pairwise correlation of biological age from samples included in both feature domains (* = Pearson correlation coefficients over R=0.5)

### FusionAge clocks illustrate the complex multimodal nature of human disease

Human diseases are not organ-specific by nature, and involve complex interplay between physiological and molecular drivers, which is captured by the multimodal architecture of FusionAge. With normalized biological age acceleration (BAA=CA-BA; transformed to z-score), FusionAge-derived clocks outperformed previously reported linear aging clocks^3,18,30^ in 24 of 30 aging-associated diseases (Fig 3b). FusionAge-DNN-ALL_EARLYFUSION outperforms all other clock implementations for predicting incident chronic pulmonary disease (AUC=0.67), hypothyroidism (AUC=0.70), other neurological disorder (AUC=0.63), rheumatoid arthritis (AUC=0.76) and solid tumor without metastasis (AUC=0.71); all FusionAge early fusion and late fusion models outperform other clocks and modality-specific clocks for predicting uncomplicated diabetes (Fig 3c), which illustrates the impact of the multimodal architecture for capturing the complexity of the disease.

**Fig. 3:**
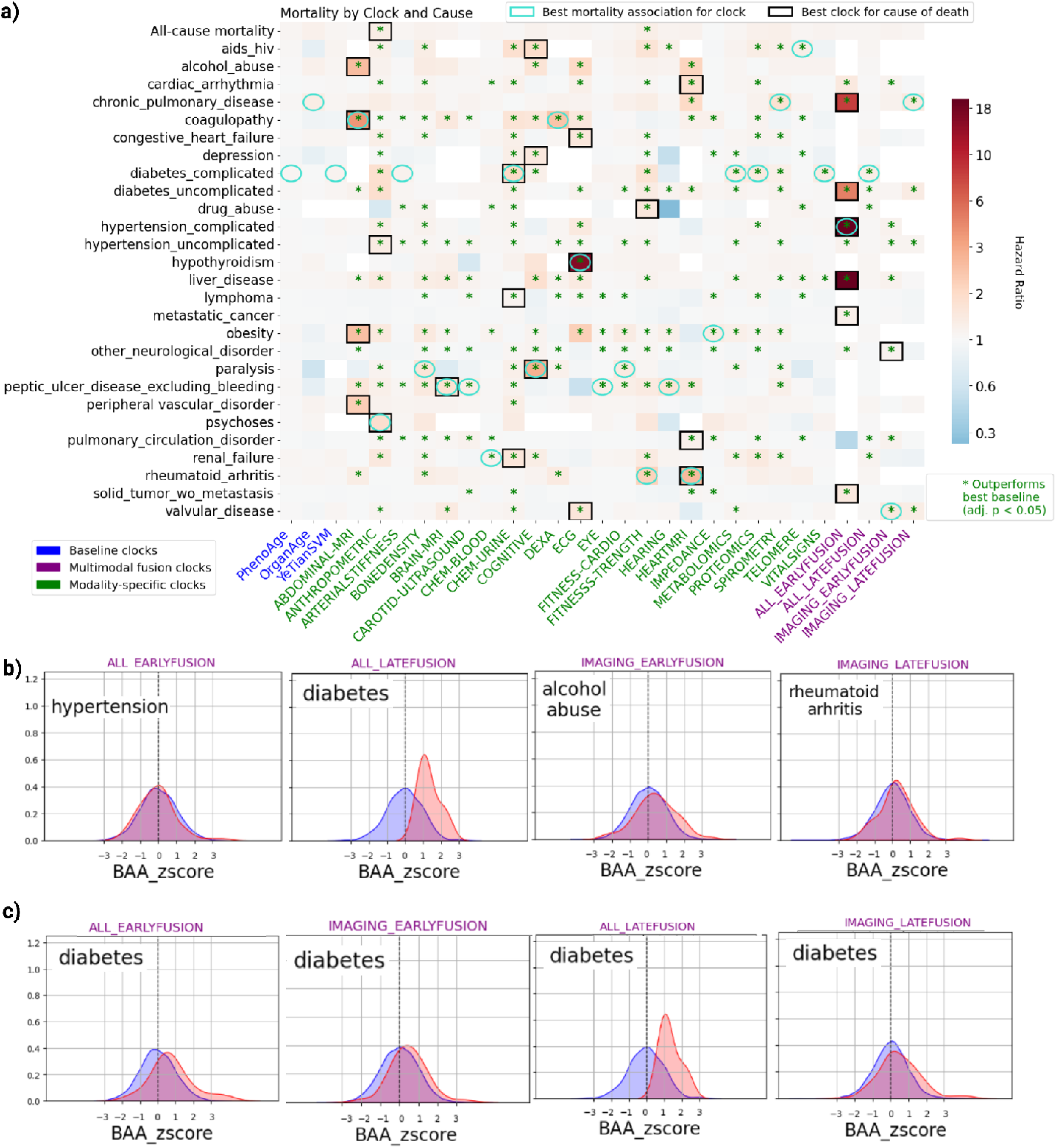
FusionAge-trained modality-specific and multimodal clocks are correlated with incident disease and mortality. a) Each cell contains the rounded log hazard ratio from Cox proportional hazards regression for testing the association between biological age acceleration and comorbidity-specific mortality. For each comorbidity, the clock that serves as the strongest predictor of mortality is outlined in bold; the clock that provides the best mortality association for a given disease is delineated with a light blue circle; green * = significant increase in predictive power of biological age acceleration for disease-specific mortality (P<0.05);b) selected distributions of biological age acceleration Z scores for the different multimodal clocks; **c)** distributions of biological age acceleration Z scores for incident diabetes, for which FusionAge-derived multimodal clocks strongly outperform modality-specific clocks

Additionally, FusionAge-DNN-IMAGING_EARLYFUSION outperforms previous clock implementations for alcohol abuse (AUC=0.99) and FusionAge-DNN-IMAGING_LATEFUSION outperforms all baselines for predicting psychoses (AUC=0.74) and weight loss related morbidities (AUC=0.65). While modality-specific clocks only include data from one modality of data collection, they often capture complex multi-organ interactions, for example shown in FusionAge-DNN-PROTEOMICS for drug abuse (AUC=0.80) and paralysis (AUC=0.65), which can exhibit subcellular perturbations in neuronal tissues across the brain and peripheral nervous system that before symptoms manifest. Additionally, the FusionAge-DNN-HEART-MRI clock is predictive for metastatic cancer, which can represent secondary malignancy in the heart (AUC=0.70).

There is a strong additive effect of biological age acceleration in incident disease association, as measured by additive prediction of disease by AUC compared to using only chronological age and sex. Most notably, the FusionAge-DNN-ALL_EARLYFUSION clock yielded strong additive performances for prediction of cardiac arrhythmia (AUC difference = 0.12), hypothyroidism (AUC difference = 0.08), and solid tumor without metastasis (AUC difference = 0.04) and the FusionAge-DNN-IMAGING_LATEFUSION clock yielded strong additive performances with respect to lymphoma (AUC difference = 0.08), other neurological disorder (AUC difference = 0.13), and valvular disease (AUC difference = 0.07).

FusionAge modality-specific and multimodal clocks outperform previous clock implementations for prediction of every category of disease-specific and all-cause mortality in Cox proportional hazards regression, controlled for chronological age and sex (Fig. 3a). The FusionAge-DNN-ANTHROPOMETRIC clock was the most predictive for all-cause mortality, as well as for uncomplicated hypertension (HR=1.3) and psychoses (HR=1.6). The urine chemistry clock was most predictive for complicated diabetes (HR=1.8). The FusionAge-DNN-COGNITIVE clock was the most predictive for mortality from paralysis, AIDS/HIV and other neurological disorders (HR=2.9, 1.7, and 1.1, respectively). The FusionAge-DNN-ABDOMINAL-MRI clock was most predictive for mortality from alcohol abuse, coagulopathy, obesity and peripheral vascular disorder. The FusionAge-DNN-BRAIN-MRI clock was most predictive for mortality from peptic ulcer disease, which could reflect cerebral driven causes of vagus nerve stimulation^38^. The FusionAge-DNN-ECG clock was most predictive for mortality from hypothyroidism (HR=15.2), congestive heart failure (HR=1.4) and valvular disease (HR=1.4). The FusionAge-DNN-HEART-MRI clock was most predictive for mortality from cardiac arrhythmia (HR=1.6) and rheumatoid arthritis (HR=2.6).

There is a strong correlation of FusionAge clocks against each other, expressed in terms of strength of mortality associations, with the strongest relationships in FusionAge-DNN-METABOLOMICS with FusionAge-DNN-ANTHROPOMETRIC (R=0.8), FusionAge-DNN-PROTEOMICS with FusionAge-DNN-CHEM-BLOOD (R=0.8), FusionAge-DNN-HEART-MRI with FusionAge-DNN-FITNESS-CARDIO (R=0.7).

### Interpretability analysis and external validation uncovers cardiorespiratory fitness as a major driver of biological age

We demonstrate the FusionAge interpretability framework on the FusionAge-ALL-EARLYFUSION clock, the clock that exhibits the strongest mortality associations across the most diseases. On the population level, summary swarm plots allow for quick interrogation of individual-level contributions of features to biological age (Fig. 4a). Aggregate score plots allow for visualization of overall negative or positive aggregate contributions of specific features to biological age (Fig. 4b). On the other hand, on the individual level, explanations for the magnitude of each individual feature’s contribution to an individual’s biological age can be quickly visualized (Fig. 4c).

**Fig. 4:**
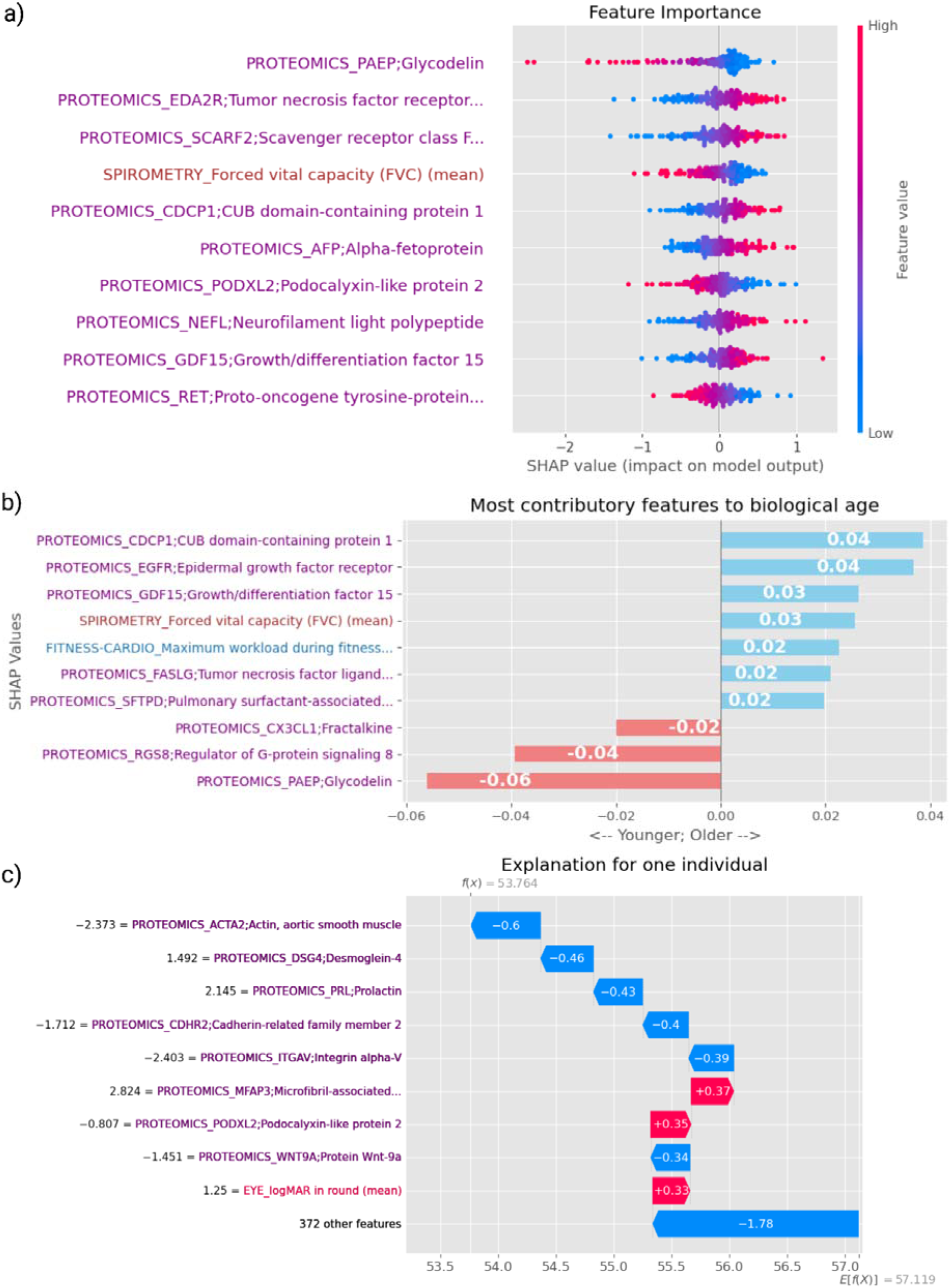
FusionAge enables both population-level and individual-level interpretability of drivers of aging. Demonstrations of the visualizations of interpretability analysis applied to the FusionAge-DNN-PROTEOMICS clock are shown: **a)** SHAP summary plot for all individuals trained on the proteomics clock. **b)** aggregate scores for each feature in the clock, which provide a quantitative objective measure for the overall relative contribution of a feature to biological age compared to other features. **c)** an individualized explanation for the drivers of biological age; features are shown in decreasing order of weight by SHAP score contribution.

The validation dataset was extracted from the US National Health and Nutrition Examination Survey (NHANES) consisting of 101,316 individuals (49,893 males and 51,423 females) with data for feature domains that overlap with those present in the training dataset (anthropometric features, vital signs, blood chemistry, urine chemistry, telomere characteristics, eye measures, cardiorespiratory fitness, skeletal muscle strength, spirometry, body impedance, DEXA scan). We trained 11 modality-specific and 2 multimodal aging clocks on the NHANES dataset; furthermore, we trained a version of the early fusion aging clock on the UKB and scored it on the NHANES dataset to predict biological age.

FusionAge retains strong performance when trained on the NHANES dataset (NHANES_EARLYFUSION, R=0.79 vs R=0.83 via FusionAge-DNN-ALL_EARLYFUSION trained on UKB), as well as when FusionAge trained on the UKB dataset is scored on the NHANES dataset (R=0.47 with DNN, R=0.57 with Lasso and ElasticNet; Fig. 5a, “EARLYFUSION_TRAIN_UKB_SCORE_NHANES”). Biological age acceleration is correlated across modality-specific and multimodal clocks. FusionAge interpretability is preserved when FusionAge modality-specific and multimodal clocks are trained on the NHANES cohort (Fig. 5c). Disease (Fig. 5b) and mortality associations are replicated in clocks trained on the NHANES cohort.

**Fig. 5:**
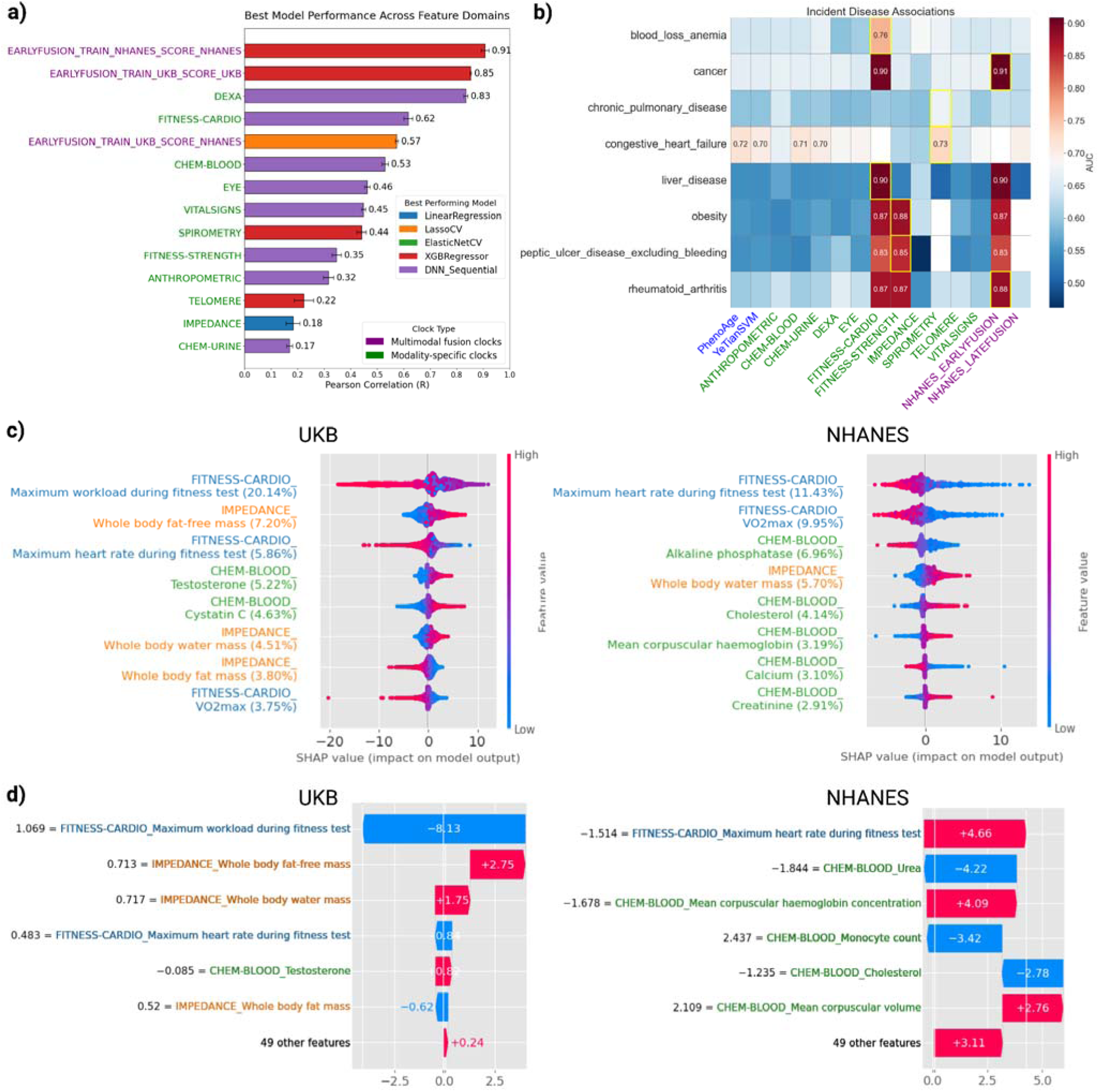
Cardiorespiratory fitness as a major driver of biological aging. A pruned multimodal early fusion aging clock using features present in both UKB and NHANES datasets is trained on both the UKB cohort (EARLYFUSION_TRAIN_UKB_SCORE_UKB) and NHANES cohort (EARLYFUSION_TRAIN_NHANES_SCORE_NHANES). Additionally, a model trained on UKB is scored on NHANES (EARLYFUSION_TRAIN_UKB_SCORE_NHANES). **a)** Pearson correlation across clocks trained in NHANES. **b)** disease associations in NHANES data. Cardiorespiratory fitness (FITNESS-CARDIO) is strongly correlated with incident disease and is the most predictive clock for blood loss anemia and liver disease. **c)** interpretability is preserved when models are trained between UKB and NHANES cohorts, which additionally validates cardiorespiratory fitness as a primary driver of biological age. **d)** example of patient-level interpretability in UKB and NHANES cohort, showing a biological age predominantly driven by cardiorespiratory fitness related features “Maximum workload during fitness test” and “VO2max” in an individual from the UKB cohort (left) and “Maximum heart rate during fitness test” and “VO2max” in an individual from the NHANES cohort.

Physical fitness is indicated as a key driver of biological aging. In both UKB and NHANES early fusion clocks, the most contributory feature to biological age was related to cardiorespiratory fitness. Crucially, the direction of this effect was consistent across both datasets: higher levels of cardiorespiratory fitness were associated with a lower biological age, while lower fitness levels were associated with a higher biological age.

In UKB, the feature “maximum workload during fitness test” was responsible for 20% of the aggregate SHAP contribution score. In NHANES, the feature “maximum heart rate during fitness test” was responsible for 11% of the aggregate SHAP contribution score. When considering the top 10 most contributory features, cardiorespiratory fitness related features are responsible for 30% of the aggregate contribution to biological age in the UKB model and 21% of the aggregate contribution to biological age in the NHANES model (Fig. 1d, 5c). Individual-level interpretability illustrates examples of strong contributions of cardiorespiratory fitness to both high and low biological age relative to chronological age (Fig. 5d). Additionally, the cardiorespiratory fitness clock FusionAge-DNN-FITNESS-CARDIO was the most predictive clock for the diseases blood loss anemia and liver disease. Another fitness-based clock, the muscle strength clock FusionAge-DNN-FITNESS-STRENGTH, was the most predictive clock for the diseases obesity and peptic ulcer disease.

### Inflammatory cytokine upregulation and tissue remodeling as putative perturbations in response to spaceflight

Finally, we trained one multimodal aging clock on a subset of the UKB cohort and scored it on a custom external dataset of four astronauts from the Inspiration4 space mission, which collected temporal information before, during and after spaceflight across multiple domains including blood proteomics, clinical chemistry markers, skin transcriptomics, and oral and nasal microbiome swabs ^33–37^. Custom FusionAge models were scored on astronauts from the Inspiration4 space mission, which was four days in duration (Fig. 1e). By FusionAge-DNN-ALL_EARLYFUSION clock (Fig. 6a), two of the four astronauts (C003 and C004) on the Inspiration4 mission exhibited increased biological age from pre-flight (L-3, 3 days before launch) to post-flight (R+1, 1 day after launch) using the data available from timepoints closest to the space mission. Interestingly the other two astronauts exhibited decreased biological age. By FusionAge-DNN-PROTEOMICS clock, three astronauts (C001, C002, C003) had increased biological age after spaceflight. On the other hand, by FusionAge-DNN-CHEM-BLOOD clock, C002 had decreased biological age immediately after spaceflight at the R+1 time point compared to immediately after spaceflight at the L-3 time point. The other 3 astronauts had increased biological age at the R+1 time point compared to L-3 time point.

**Fig. 6:**
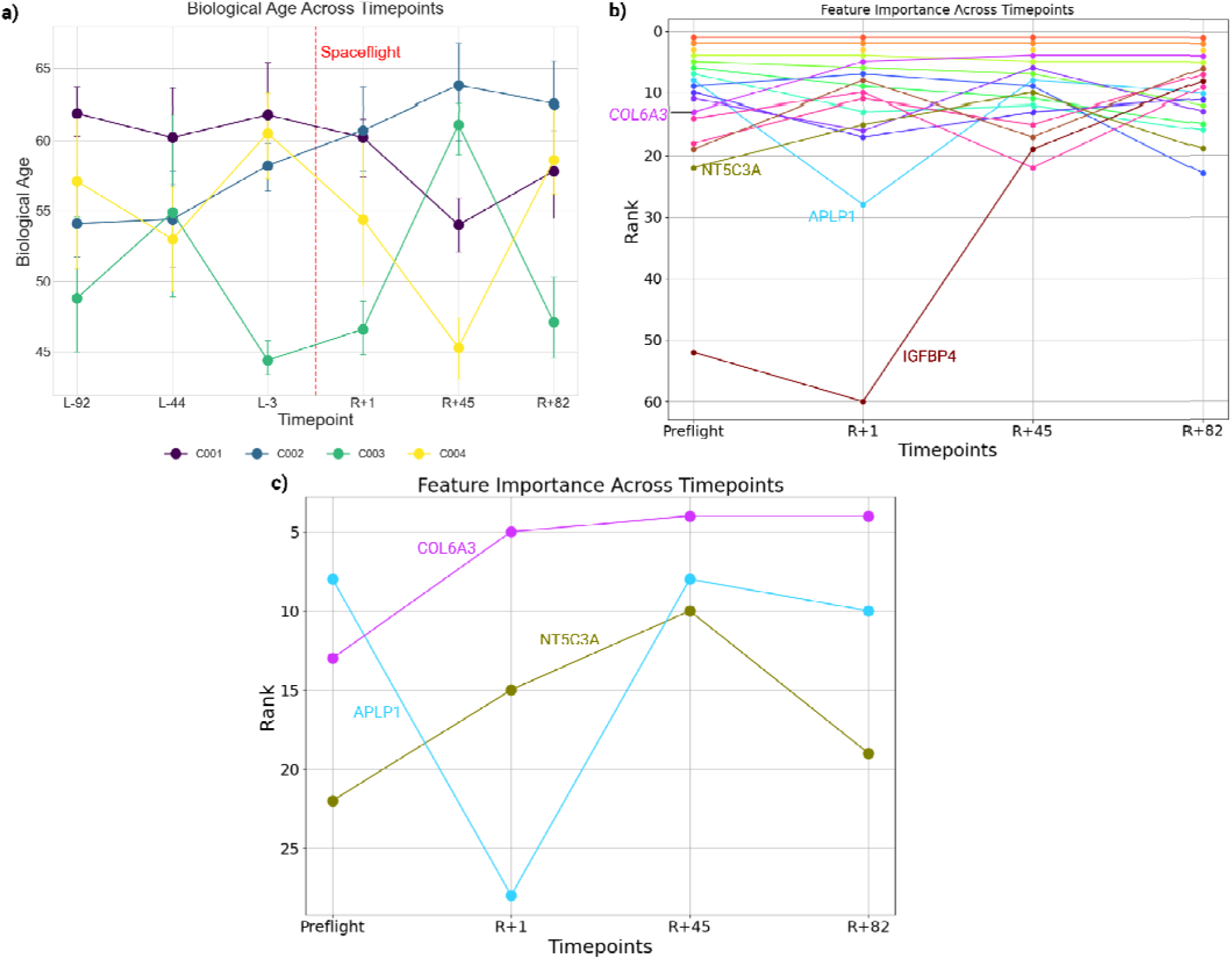
Drivers of biological age change in response to spaceflight in astronauts. **a)** multimodal CHEM-BLOOD and PROTEOMICS clock of the 4 crew members of the Inspiration4 mission, across timepoints before and after the mission, biological age shown as mean ± SEM; **b)** change in ranking of feature importance across longitudinal timepoints (preflight, R+1, R+45, R+82) for the union of the top 10 most important features across all timepoints; **c)** change in ranking of feature importance across longitudinal timepoints (preflight, R+1, R+45, R+82) for the features NT5C3A, APLP1, and COL6A3 from the PROTEOMICS domain.

With the FusionAge-DNN-ALL_EARLYFUSION clock, when comparing R+1 and R+45, two astronauts (C001 and C004) decreased their biological age, which may reflect longer term recovery. Of note, both C001 and C004 are male while C002 and C003 are female, and this may reflect sex-specific differences in adaptations to recovery after spaceflight.

By interpretability analysis, the features in the FusionAge-DNN-ALL_EARLYFUSION clock that contribute most to higher biological age in all preflight and post-flight timepoints include the proteomics features LTBP2, GDF15 and EFEMP1, which likely represent general baseline drivers of biological age that are not significantly related to spaceflight. The most contributory feature to lower biological age across all timepoints is the protein COL6A3. Given the aforementioned proteins’ high contribution across both pre-flight and post-flight timepoints, they can be considered general proteins associated with aging that are minimally influenced by spaceflight.

The protein NT5C3A exhibits a 41% increase in contribution to increased biological age at the immediate post-flight timepoint compared to pre-flight timepoints (aggregate contribution score 16.65 vs. average 11.8 preflight). Interestingly, there is a reversion of the contribution towards pre-flight levels at the R+45 and R+82 timepoints (aggregate contribution scores 8.29 and 10.30, respectively). Previous literature showed that NT5C3A has been found to play a role in suppressing cytokine production and inhibiting the NF-κB pathway, which is involved in immune and inflammatory responses. Furthermore, due to the known increase in radiation during spaceflight, the increase in NT5C3A after spaceflight could be expected due to the known involvement of the protein in nucleotide catabolism and associated DNA repair processes ^39,40^ (Fig. 6b-c).

The protein, APLP1, expressed in cardiomyocytes, exhibited a 35% decreased contribution to biological age at the R+1 time point compared to preflight (aggregate contribution score −1.48 vs average - 2.28). The contribution returned to baseline levels at the R+45 timepoint (aggregate contribution score 2.57) (Fig. 6b-c). While limited accounts of APLP1 changes have been reported in the context of spaceflight, the finding may be related to upregulation of oxidative stress, or changes in expression of cell cycle arrest and apoptosis regulatory genes, including upregulation of Cdkn1a (p21)^41^.

The protein, COL6A3 (collagen type VI alpha 3 chain), exhibits significantly increased contribution to biological age at the R+1 time point compared to preflight (aggregate contribution score −5.29 vs average - 0.24; feature importance ranking 5 at R+1 postflight vs ranking 16 preflight) and remains as a top driver of biological aging at the subsequent post-flight R+45 and R+82 time points (SHAP rank 4) (Fig. 6b-c). The changes in COL6A3 in response to spaceflight have been previously characterized in skin (proposed to be involved in extracellular matrix remodeling after spaceflight ^42,43^) and muscle (increased expression after spaceflight^44^) and has been proposed to play a continued role in readaptation upon return to earth after spaceflight, which may reflect the continued importance of this protein in driving the biological age of astronauts in all post-flight timepoints.

## Discussion

In this study, we describe a framework for biological age estimation that demonstrates two key advances: first, that multimodal data integration provides a more robust estimation of biological age than any single data type, and second, that nonlinear models such as deep neural networks consistently outperform linear approaches. Our comprehensive multimodal clock, FusionAge-DNN-ALL_LATEFUSION, achieved a Pearson correlation of R=0.95 with chronological age, substantially higher than baseline linear models like PhenoAge^3^ (R=0.75) and other clocks from the literature that also use nonlinear methods (Sagers et al.^19^ random forest (R=0.71), Aging.AI^19,20^, deep neural network (R=0.87), Rahman et al ^21^, deep neural network (R=0.8))

FusionAge-derived biological age is strongly associated with incident disease and aging-related mortality. Furthermore, since FusionAge-derived aging clocks are based on modalities rather than predetermined features related to specific organ systems, modality-specific clocks often exhibit stronger disease or mortality associations than other implementations of aging clocks or FusionAge multimodal clocks. Here, we showed that compared with previously reported linear aging clocks, the FusionAge-derived biological age acceleration is more predictive of incident disease in 24 of 30 aging-associated diseases. For example, FusionAge clocks outperform previously reported clock implementations in diseases such as fluid and electrolyte disorders (FusionAge-DNN_ABDOMINAL-MRI clock), obesity (FusionAge-DNN_ABDOMINAL-MRI clock), hypothyroidism (FusionAge-DNN_CAROTID-ULTRASOUND clock), and psychoses (FusionAge-DNN_METABOLOMICS clock). Multimodal clocks were useful for various diseases which have multi-organ sequelae (e.g., alcohol abuse, cardiac arrhythmia, coagulopathy, diabetes, hypertension, liver disease, metastatic cancer, paralysis).

Similar trends are noted for mortality associations. For all diseases, either a FusionAge modality-specific clock or a multimodal FusionAge clock outperformed other previous implementations (PhenoAge, OrganAge, YeTianSVM). The strongest mortality associations are observed via the FusionAge-DNN-ECG clock for hypothyroidism, and the FusionAge-DNN-ALL_EARLYFUSION clock for complicated hypertension, liver disease, chronic pulmonary disease and diabetes. Strong disease and mortality associations can facilitate future efforts to use multimodal aging clocks for applications such as patient stratification in healthcare settings or endpoints for interventional studies involving putative geroprotective therapeutics. An important future direction will be to apply this framework to investigate the impact of specific interventions on biological aging. For example, the effect of drugs with geroprotective potential, such as metformin, or specific lifestyle changes could be quantified by tracking changes in multimodal biological age over time. This would provide a powerful tool for evaluating strategies aimed at promoting healthier aging.

From external validation analysis, we demonstrate the retained performance when analogous versions of the FusionAge early fusion clocks were trained on both the UKB (R=0.83) and NHANES datasets (R=0.88) and scored respectively. When the clock was trained on the UKB and subsequently scored on NHANES, biological age retained its correlation with chronological age (R=0.57) albeit with a drop-off in performance, which may be due to intrinsic differences in the populations of individuals in the two datasets, which were sampled from the United Kingdom and the United States respectively. Furthermore, a majority of individuals from the UKB are from an older population (mean age 56.8, range 39 to 70) while the ages of individuals from the same age range in NHANES were more uniformly distributed (mean age 43.4). Nonetheless, we demonstrate the potential for portability of the FusionAge approach that can be refined in future work.

While interpretability remains a challenge for adoption of aging clocks in a clinical environment, we demonstrate the usage of the FusionAge interpretability module in understanding the top drivers of biological age at the aggregate cohort level as well as the individual level. In both the UKB and NHANES datasets, the top drivers of biological age are dominated by features related to cardiorespiratory fitness (30% of the aggregate contributions in UK Biobank, 21% of aggregate contributions in NHANES) (Fig. 5c). This finding forms a strong basis for future hypothesis generation for mechanistic studies that may involve interactions between cardiorespiratory fitness and other features (e.g., proteins, metabolites).

A version of the FusionAge multimodal clock on a cohort of four astronauts’ preflight and postflight features surrounding the Inspiration4 space mission, which serves as a proof-of-concept that lays a foundation for future work focusing on biological age estimation in larger cohorts of astronauts and other populations of individuals undergoing extreme physical stressors. For such individuals, longitudinal profiling of biological age can be useful for tracking their adaptation to spaceflight and subsequent return, especially using domains of information readily obtainable from healthcare settings (e.g., clinical chemistry tests requiring blood draws) or personal home settings (e.g., fitness tests).

Our work should motivate future efforts to train multimodal aging clocks that incorporate multiple data domains including biospecimens, physical measures, imaging and functional tests. While not all domains may be present in all situations, our framework can be successfully ported to other populations. We show that spaceflight induces various biological aging changes, and further interrogation of the specific features driving the changes can provide actionable insights on devising effective countermeasures in preparation.

In summary, the FusionAge framework enables robust biological age estimation by integrating diverse data types with nonlinear models. While our demonstration focused on clinical, -omics, and functional data, the framework is highly customizable for future work incorporating other sources like epigenetics or the microbiome ^45^. We validate the impact of cardiorespiratory fitness as a major, cross-cohort driver of biological age and demonstrate the framework’s utility in a novel context like spaceflight, informing future mechanistic studies and applications in human health.

## Methods

### General overview of FusionAge framework

Data processing and cohort construction are performed to create feature sets for construction of modality-specific clocks and multimodal clocks. Validation included assessment of performance metrics, incident disease and mortality associations across aging-related diseases, interpretability analysis of the major drivers of biological age estimation. Finally, a deep learning based multimodal early fusion model was trained and scored on a cohort of astronauts to demonstrate the application of FusionAge (Fig. 1).

### Data

The data source used for training the aging clocks was the UK Biobank, a roughly 500,000 patient biomedical database and research resource which includes clinical, biological, genetic, and metabolomic data. The data was taken from UK Biobank application 47137 and all participants consented to be subjects in the study. At time of recruitment, patient ages ranged from 37 to 73 years. For all patients, all biospecimen-based testing and physical assessments were performed at an initial baseline assessment visit, which spanned from 2006 to 2010. A subset of these had subsequent visits after the initial assessment.

Approximately 20,000 participants had a first repeat assessment (2012-2013), approximately 85,000 had a first imaging visit (from 2014 to ongoing), and approximately 9,000 had a second imaging visit (from 2019 to ongoing). Details on available phenotypes can be found online (https://biobank.ndph.ox.ac.uk/showcase/) and all participants provided informed consent.

The validation dataset was a cohort extracted from US National Health and Nutrition Examination Surveys (NHANES) (https://wwwn.cdc.gov/nchs/nhanes/Default.aspx), which included approximately 95,000 individuals sampled between 1999-2017.

The use case dataset was a cohort of four individuals from the Inspiration4 space mission, which collected temporal information before, during and after spaceflight across multiple domains including blood proteomics, metabolomics, and chemistry markers, skin transcriptomics, and oral and nasal microbiome swabs ^33–37^. Samples were collected from the crew members at specified time points relative to the spaceflight mission: L-92, L-3, L-44, R+1, R+45, R+82, R+194, representing the number of days before launch date (“L”) or after the return to earth date (“R”).

### Data Processing

#### Data extraction and cohort construction

From the UK Biobank, feature sets for each modality of data collection were extracted (Fig. 1a). Modality-specific clocks used features from the following domains anthropometric features, arterial stiffness, bone density, blood chemistry, urine chemistry, cognitive test, eye measures, cardiorespiratory fitness test, strength test, hearing test, body impedance, metabolomics, proteomics, spirometry test, telomeres, vital signs, abdominal MRI, brain MRI, carotid ultrasound, DEXA scan, electrocardiogram and heart MRI. Multimodal clocks were trained from two subsets of modalities: one using information from the baseline visit (anthropometric features, arterial stiffness, bone density, blood chemistry, urine chemistry, cognitive test, eye measures, cardiorespiratory fitness test, strength test, hearing test, body impedance, metabolomics, proteomics, spirometry test, telomeres and vital signs), and one from the second follow-up visit which served as the first imaging visit (abdominal MRI, brain MRI, carotid ultrasound, DEXA scan, electrocardiogram and heart MRI) (Fig. 1a).

For each modality-specific clock, individuals were included if they had data recorded for any of the features involved in that modality. For each multimodal clock, individuals were included if they had data recorded for all of the modalities involved in the clock. Across all clocks trained, individuals were included in the cohort for that clock only if they had a documented age at the time of the visit.

### Aging Clock Construction

#### Problem formulation

Individual (modality-specific clocks) and combined (multimodal clocks) featured domains are used as predictive features in a regression problem for predicting chronological age (CA, age at time of visit). Multimodal clocks are formulated with both early fusion and late fusion architectures. Subsequently, the following regression algorithms are trained: linear regression (LR), lasso, elastic net, XGBoost, and deep neural network (DNN). The predicted age for an individual is the biological age (BA). The biological age acceleration is the difference between biological age and chronological age (BAA=BA-CA). From here on, biological age derived from a specific algorithm ALGO and modality MODALITY is referred to as FusionAge-ALGO-MODALITY (e.g., FusionAge-LR-CHEM-BLOOD, FusionAge-DNN-CHEM-BLOOD). Biological age derived from a multimodal feature set will be referenced by a label for that feature set (e.g., FusionAge-DNN-ALL_LATEFUSION, FusionAge-DNN-ALL_EARLYFUSION) (Table 1 lists all clocks and naming conventions).

Features from each domain are constructed in the following manner. For continuous features (e.g., plasma hemoglobin A1c level), the raw value is used. If a feature was tested in both the left and right side of the body (e.g., handgrip strength, arm fat mass, heel bone mineral density), then the mean of the readings on each side of the body was used, with the following exceptions: for the strength test, individual values on each side in addition to mean values were used, in order to capture gross muscular strength imbalances. Missing values were imputed with the median value for that feature across all individuals. After imputation each feature vector was standardized.

If a feature was tested more than one time at the visit (e.g., blood pressure, forced expiratory volume in the first second (FEV1)), the mean of all readings was used. For the spirometry test, additional features were constructed for the trend in values for forced vital capacity (FVC) and forced expiratory volume in 1-second (FEV1) across all readings, in order to account for differing changes in fatigue over time between individuals.

For the cardiorespiratory fitness test, the maximal oxygen consumption (VO2max) was constructed with the heart rate and workload waveforms from the fitness test, according to the American College of Sports Medicine guidelines ^46^.

#### Regression to model biological age

For each clock, the following regression algorithms were trained: linear regression, lasso regression, elastic net regression, XGBoost, deep neural network. The training process for each algorithm was performed as follows. The combined feature matrix was split into a training set (70%), cross validation set (10%) and test set (20%).

Feature selection was performed using recursive feature elimination via linear regression. Model selection was conducted via 10-fold cross validation on the cross validation set in order to tune the parameters for that specific algorithm (e.g., values for alpha in lasso, learning rate and max tree depth for XGBoost). Using the best model based on Pearson correlation coefficient (R) from the model selection process, 10-fold cross validation was performed on the training set in order to determine model weights for each feature. The model from the fold with the best performance by Pearson correlation coefficient was used to score the test set. Model construction was performed with the Python packages: scikit-learn version 1.0.2 (linear regression, lasso, elastic net) ^47^, xgboost version 1.6.2 (XGBoost)^48^, and PyTorch version 1.13.1 ^49^.

### Performance Analysis

#### Performance Metrics

Trained models were scored on the test set via Pearson correlation coefficient (R), mean absolute error (MAE) and root mean squared error (RMSE). Performance of the final FusionAge models were compared against biological age obtained from linear models (PhenoAge^3^, OrganAge^18^, YeTianSVM^30^).

#### Model Interpretability

We derive interpretations of feature contributions to biological age via the Shapley additive explanations (SHAP) method, providing an assessment of the additive contribution of individual features to the prediction of biological age on a local level^50^. To determine the impact of specific features on the overall biological age, an aggregated SHAP score was devised as follows:

For each individual i and each feature j, the contribution score c is defined as

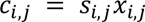

The aggregate contribution of the feature j is

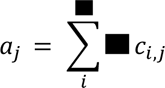

A rank ordered list of all *α_j_* represents the ranking of features from most contributory to least contributory.

#### Disease and mortality associations

Using the multimodal modal with the strongest correlation coefficient, FusionAge-DNN-ALL_EARLYFUSION, we computed associations of FusionAge-DNN with incident diseases of aging via binary classification with logistic regression using biological age acceleration (BAA = CA-BA), chronological age (CA) and sex as features, assessed by area under the receiver operating curve (AUC). The additional predictive effect of FusionAge-derived biological age acceleration was compared with chronological age alone. To compute the additive effect, the AUC obtained from using BAA, CA and sex as features (*AUC_with BAA_*)was compared against the AUC obtained from using only CA and sex as features (*AUC_without BAA_*), i.e., *AUC_additional_* = *AUC_with BAA_* - *AUC_without BAA_*.

Associations of FusionAge-DNN-ALL_EARLYFUSION normalized biological age acceleration (BAA = CA-BA) with all-cause mortality, as well as mortality related to aging-related diseases, were performed via the Cox proportional hazards model, whereby hazard ratios for mortality were computed for BA, controlled for chronological age ^51^. Diseases were extracted and grouped into categories via the Python icd package ^52^.

### External validation

#### Portability to external datasets

To assess the portability of the FusionAge framework to external data sources, for the NHANES external validation dataset, a pruned version of the FusionAge-DNN-ALL_EARLYFUSION clock was trained using the features that are present in both the UK Biobank and the external NHANES validation dataset: blood chemistry, urine chemistry, eye measures, cardiorespiratory fitness, body impedance (Fig. 1d). Additionally, modality-specific clocks are trained for these feature sets. Disease and mortality associations are computed using normalized biological age acceleration for all clocks in both datasets. For the multimodal early fusion clock, the pruned UK Biobank clock was also scored on the NHANES dataset (results reported in Fig. 5a, referenced as EARLYFUSION_TRAIN_UKB_SCORE_UKB (UK Biobank clock scored on UK Biobank data), EARLYFUSION_TRAIN_NHANES_SCORE_NHANES (NHANES clock scored on NHANES data), EARLYFUSION_TRAIN_UKB_SCORE_NHANES (UK Biobank clock scored on NHANES data)). In both UK Biobank and NHANES, feature rankings describing aggregate contributions to biological age are computed in the FusionAge-DNN-ALL_EARLYFUSION clocks trained on the respective cohort. In order to rank modalities that drive contribution to biological age, the top 10 most contributory individual features for biological age are grouped by modality and the proportion of aggregate contribution for that modality is determined.

### Use case demonstration: spaceflight-induced biological age changes

We trained a multimodal model using data modalities in common between the UK Biobank dataset and the Inspiration4 dataset (Fig. 1e). We trained modality-specific clocks with blood chemistry, metabolomics and proteomics modalities, as well as a multimodal clock with all of these modalities.

## Data Availability

All datasets used are publicly available online at https://wwwn.cdc.gov/nchs/nhanes/Default.aspx (NHANES), https://www.ukbiobank.ac.uk (UK Biobank), https://osdr.nasa.gov/bio/repo/data/missions/SpaceX%20Inspiration4 (Inspiration 4 space mission).

## Code Availability

The code for FusionAge as well as code to conduct the supporting analyses are available as zipped archives, provided in the Supplementary Information.

## Notes

### Competing Interest Statement

The authors have declared no competing interest.

### Funding Statement

This study did not receive any funding.

## References

1. Hannum, G. et al. Genome-wide methylation profiles reveal quantitative views of human aging rates. Mol. Cell 49, 359–367 (2013).

2. Horvath, S. DNA methylation age of human tissues and cell types. Genome Biol. 14, R115 (2013).

3. Levine, M. E. et al. An epigenetic biomarker of aging for lifespan and healthspan. Aging 10, 573–591 (2018).

4. Higgins-Chen, A. T. et al. A computational solution for bolstering reliability of epigenetic clocks: Implications for clinical trials and longitudinal tracking. Nat Aging 2, 644–661 (2022).

5. Belsky, D. W. et al. DunedinPACE, a DNA methylation biomarker of the pace of aging. Elife 11, (2022).

6. Lu, A. T. et al. DNA methylation GrimAge strongly predicts lifespan and healthspan. Aging 11, 303–327 (2019).

7. Klemera, P. & Doubal, S. A new approach to the concept and computation of biological age. Mech. Ageing Dev. 127, 240–248 (2006).

8. Belsky, D. W. et al. Quantification of the pace of biological aging in humans through a blood test, the DunedinPoAm DNA methylation algorithm. Elife 9, (2020).

9. Belsky, D. W. et al. Quantification of biological aging in young adults. Proc. Natl. Acad. Sci. U. S. A. 112, E4104–10 (2015).

10. Macdonald-Dunlop, E. et al. A catalogue of omics biological ageing clocks reveals substantial commonality and associations with disease risk. Aging 14, 623–659 (2022).

11. Rutledge, J., Oh, H. & Wyss-Coray, T. Measuring biological age using omics data. Nat. Rev. Genet. 23, 715–727 (2022).

12. Robinson, O. & Lau, C.-H. E. Measuring biological age using metabolomics. Aging 12, 22352–22353 (2020).

13. Hertel, J. et al. Measuring Biological Age via Metabonomics: The Metabolic Age Score. J. Proteome Res. 15, 400–410 (2016).

14. Lassen, J. K. et al. Large-Scale metabolomics: Predicting biological age using 10,133 routine untargeted LC-MS measurements. Aging Cell 22, e13813 (2023).

15. Robinson, O. et al. Determinants of accelerated metabolomic and epigenetic aging in a UK cohort. Aging Cell 19, e13149 (2020).

16. Buergel, T. et al. Metabolomic profiles predict individual multidisease outcomes. Nat. Med. 28, 2309–2320 (2022).

17. Oh, H. S.-H. et al. Plasma proteomics in the UK Biobank reveals youthful brains and immune systems promote healthspan and longevity. bioRxiv 2024.06.07.597771 (2024) doi:10.1101/2024.06.07.597771.

18. Oh, H. S.-H. et al. Organ aging signatures in the plasma proteome track health and disease. Nature 624, 164–172 (2023).

19. Sagers, L., Melas-Kyriazi, L., Patel, C. J. & Manrai, A. K. Prediction of chronological and biological age from laboratory data. Aging 12, 7626–7638 (2020).

20. Putin, E. et al. Deep biomarkers of human aging: Application of deep neural networks to biomarker development. Aging 8, 1021–1033 (2016).

21. Ashiqur Rahman, S., et al. Deep learning for biological age estimation. Brief. Bioinform. 22, 1767–1781 (2021).

22. Acosta, J. N., Falcone, G. J., Rajpurkar, P. & Topol, E. J. Multimodal biomedical AI. Nat. Med. 28, 1773–1784 (2022).

23. Huang, S.-C., Pareek, A., Seyyedi, S., Banerjee, I. & Lungren, M. P. Fusion of medical imaging and electronic health records using deep learning: a systematic review and implementation guidelines. NPJ Digit Med 3, 136 (2020).

24. Moon, K. R. et al. Visualizing structure and transitions in high-dimensional biological data. Nat. Biotechnol. 37, 1482–1492 (2019).

25. Kuchroo, M. et al. Multiscale PHATE identifies multimodal signatures of COVID-19. Nat. Biotechnol. 40, 681–691 (2022).

26. Wang, T. et al. MOGONET integrates multi-omics data using graph convolutional networks allowing patient classification and biomarker identification. Nat. Commun. 12, 3445 (2021).

27. Kwolek, B. & Kepski, M. Human fall detection on embedded platform using depth maps and wireless accelerometer. Comput. Methods Programs Biomed. 117, 489–501 (2014).

28. Masison, J. et al. A modular computational framework for medical digital twins. Proc. Natl. Acad. Sci. U. S. A. 118, (2021).

29. Fisher, C. K., Smith, A. M., Walsh, J. R., Coalition Against Major Diseases & Abbott, Alliance for Aging Research, Alzheimer’s Association, Alzheimer’s Foundation of America, AstraZeneca Pharmaceuticals LP, Bristol-Myers Squibb Company, Critical Path Institute, CHDI Foundation, Inc., Eli Lilly and Company, F. Hoffmann-La Roche Ltd, Forest Research Institute, Genentech, Inc., GlaxoSmithKline, Johnson & Johnson, National Health Council, Novartis Pharmaceuticals Corporation, Parkinson’s Action Network, Parkinson’s Disease Foundation, Pfizer, Inc., sanofi-aventis. Collaborating Organizations: Clinical Data Interchange Standards Consortium (CDISC), Ephibian, Metrum Institute. Machine learning for comprehensive forecasting of Alzheimer’s Disease progression. Sci. Rep. 9, 13622 (2019).

30. Tian, Y. E. et al. Heterogeneous aging across multiple organ systems and prediction of chronic disease and mortality. Nat. Med. 29, 1221–1231 (2023).

31. Mandsager, K. et al. Association of Cardiorespiratory Fitness With Long-term Mortality Among Adults Undergoing Exercise Treadmill Testing. JAMA Netw Open 1, e183605 (2018).

32. for Disease Control, C. & (cdc), P. National health and nutrition examination survey data. Hyattsville, MD: US Department of Health and Human Services, Centers for Disease Control and Prevention 2020, (2010).

33. Overbey, E. G. et al. The Space Omics and Medical Atlas (SOMA) and international astronaut biobank. Nature (2024) doi:10.1038/s41586-024-07639-y.

34. Overbey, E. G. et al. Collection of biospecimens from the inspiration4 mission establishes the standards for the space omics and medical atlas (SOMA). Nat. Commun. 15, 4964 (2024).

35. Kim, J. et al. Single-cell multi-ome and immune profiles of the Inspiration4 crew reveal conserved, cell-type, and sex-specific responses to spaceflight. Nat. Commun. 15, 4954 (2024).

36. Sanders, L. M. et al. Inspiration4 data access through the NASA Open Science Data Repository. NPJ Microgravity 10, 56 (2024).

37. Jones, C. W. et al. Molecular and physiologic changes in the SpaceX Inspiration4 civilian crew. Nature (2024) doi:10.1038/s41586-024-07648-x.

38. Stevens, H., Guin, G. H. & Gilbert, E. F. GASTROINTESTINAL ULCERATION AND CENTRAL NERVOUS SYSTEM LESIONS. Am. J. Dis. Child. 106, 613–619 (1963).

39. Kashirina, D. N. et al. The molecular mechanisms driving physiological changes after long duration space flights revealed by quantitative analysis of human blood proteins. BMC Med Genomics 12, 45 (2019).

40. Al-Haj, L. & Khabar, K. S. A. The intracellular pyrimidine 5’-nucleotidase NT5C3A is a negative epigenetic factor in interferon and cytokine signaling. Sci Signal 11, (2018).

41. Kumar, A., Tahimic, C. G. T., Almeida, E. A. C. & Globus, R. K. Spaceflight Modulates the Expression of Key Oxidative Stress and Cell Cycle Related Genes in Heart. Int J Mol Sci 22, (2021).

42. Cope, H. et al. More than a Feeling: Dermatological Changes Impacted by Spaceflight. Res Sq (2023) doi:10.21203/rs.3.rs-2367727/v1.

43. Cope, H. et al. Transcriptomics analysis reveals molecular alterations underpinning spaceflight dermatology. Commun Med (Lond) 4, 106 (2024).

44. Blottner, D. et al. Space Omics and Tissue Response in Astronaut Skeletal Muscle after Short and Long Duration Missions. Int J Mol Sci 24, (2023).

45. Jokai, M. et al. DNA methylation clock DNAmFitAge shows regular exercise is associated with slower aging and systemic adaptation. Geroscience (2023) doi:10.1007/s11357-023-00826-1.

46. ) S. G. (ph, Dwyer, G. B. & American College of Sports Medicine. ACSM’s Metabolic Calculations Handbook. (Lippincott Williams & Wilkins, 2007).

47. Pedregosa, F. et al. Scikit-learn: Machine Learning in Python. J. Mach. Learn. Res. abs/1201.0490, (2011).

48. Chen, T. & Guestrin, C. XGBoost: A Scalable Tree Boosting System. in Proceedings of the 22nd ACM SIGKDD International Conference on Knowledge Discovery and Data Mining 785–794 (Association for Computing Machinery, New York, NY, USA, 2016).

49. Paszke, A., et al. PyTorch: An imperative style, high-performance deep learning library. Adv. Neural Inf. Process. Syst. abs/1912.01703, (2019).

50. Lundberg, S. M. & Lee, S.-I. A unified approach to interpreting model predictions. Adv. Neural Inf. Process. Syst. 4765–4774 (2017).

51. Cox, D. R. Regression models and life-tables. J. R. Stat. Soc. 34, 187–202 (1972).

52. icd 0.1.3. PyPI https://pypi.org/project/icd/.

